# Oxygen saturation instability in suspected covid-19 patients; contrasting effects of reduced V_A_/Q and shunt

**DOI:** 10.1101/2020.12.17.20248126

**Authors:** J.Gareth Jones

**Author notes:** Woodlands, Rufforth, York, YO23 4QF, UK.

## Abstract

Patients in the UK at risk of Covid-19 pneumonia, but not needing immediate hospital attention, are to be given pulse oximeters to identify a fall in oxygen saturation (SaO2 or SpO2) at home. A recent finding in Covid-19 pneumonia is a dominant reduction in ventilation to perfused alveoli (V_A_/Q). A mathematical model of gas exchange was used to predict the effect of shunt or reduced V_A_/Q on SaO2 stability inferred from the slope of the PIO2 vs SaO2 curve as it intersected the line representing ambient PIO2. A ±1 kPa variation in PIO2 predicted a 1.5% and 8% change in SpO2 with 15% shunt and 0.4 V_A_/Q respectively. As a consistency check, two patients with pre-existing lung disease and 12 hour continuous SpO2 monitoring breathing air had gas exchange impairment analysed in terms of shunt and reduced V_A_/Q. The patient with 16% shunt and normal V_A_/Q had a stable but reduced SpO2 (circa 93±1%) throughout the 12 hr period. The patient with a V_A_/Q reduced to 0.48 had SpO2 ranging from 75-95% during the same period. SpO2 monitoring in suspected covid-19 patients should focus on SpO2 varying >5% in 30 minutes. Such instability in at risk patients is not diagnostic of Covid-19 pneumonia but this may be suspected from a dominant reduction in V_A_/Q if episodic hypoxaemia has progressed from a stable SpO2.

## INTRODUCTION

200,000 patients in England at risk of complications with Covid-19, but not needing immediate hospitalisation, are to be given pulse oximeters to identify deterioration in oxygen saturation (SpO2) at home,[1]. This is to create “virtual Covid wards” of at risk patients who take SpO2 readings and relate these to their health teams. Mortality risk increases and admission to hospital is indicated if SpO2 falls to 94% or less,[1].

A new finding may transform thinking about SpO2 monitoring in incipient Covid-19 Adult Respiratory Distress Syndrome (ARDS). This is a dissociation between severe hypoxemia and well-preserved alveolar gas volume in Covid-19 ARDS causing reduced ventilation to perfused alveoli (V_A_/Q) with high compliance and radiological sparing. This is virtually never seen in most forms of ARDS where large shunt, extensive lung radio-opacities and low compliance dominate,[2]. A qualitative relationship has been reported between reduced V_A_/Q in patients with pre-existing lung disease and unstable SpO2 measured during 12 hour periods of continuous monitoring,[3,4].

A model of pulmonary gas exchange is used here to examine the effect of a reduction in V_A_/Q or increase in shunt on the position and slope of the PIO2 vs SpO2 curve as it intersects the line representing 21 kPa PIO2, i.e breathing air at sea level. From this will be inferred the stability of SaO2 with changes in distribution of ventilation or shunt. Newly available methods have enabled more precise discrimination of shunt and reduced VA/Q to examine the relationship between these entities and SpO2 stability from previous studies,[5,6]. It is proposed that SpO2 instability is a sign of reduced V_A_/Q which may be a diagnostic feature of incipient Covid-19 ARDS.

## BACKGROUND

V_A_/Q and shunt can be derived non-invasively using a computer algorithm to relate SpO2 to inspired oxygen pressure (PIO2) in a PIO2 vs SpO2 diagram,[4-7]. An unstable SpO2 is explained by rightward shift of the PIO2 vs SpO2 relationship when V_A_/Q and shunt change and is shown diagrammatically in Fig 1. The shape of the oxygen dissociation curve (ODC, red line, i.e. PaO2 vs SaO2,) shifted to the right by PaCO2/R (blue arrow), provides a PIO2 vs SpO2 curve for the normal lung (blue line). Reducing V_A_/Q shifts this curve further to the right (green arrow and line) and as it crosses the 21kPa PIO2 line, representing air breathing, its gradient becomes increasingly steep. Small changes in V_A_/Q (or PIO2) cause large changes in SpO_2_. Increasing shunt bends the curve downwards,[4-6] so that large changes in PIO_2_ cause small changes in SpO_2_ emphasising the unreliability of the P_a_O_2_/F_I_O_2_ as an index of oxygen exchange,[8].

**Fig 1.**
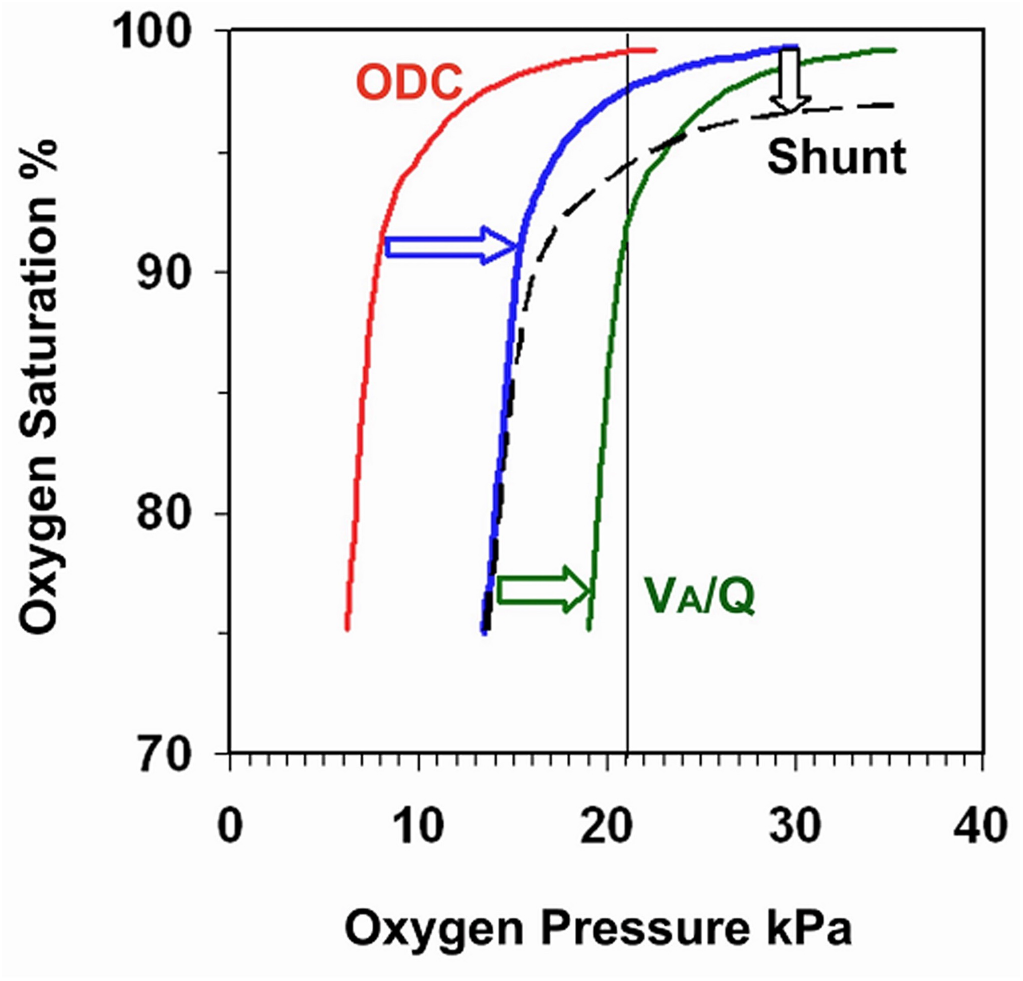
Shows how the Oxygen Dissociation Curve (ODC) determines the shape of the normal lung PIO2 vs SPO2 curve (blue) and the effects of reducing VA/Q (green) or increasing shunt (black). Vertical line is at 21 kPa PIO2.

An algorithm based on a three compartment lung model was recently developed to analyse the slope of the PIO2 vs SpO2 curve in pre term infants with broncho pulmonary dysplasia,[5-7]. This showed that slope was greatest in those with a homogeneous reduction of V_A_/Q to circa 0.4. Slope was considerably less with inhomogeneous pulmonary disease with or without increased shunt.

## METHODS

The effects of reducing V_A_/Q or increasing shunt on PIO2 vs SpO2 curves were derived using a mathematical model of pulmonary gas exchange described by Olszowska and Wagner,[9]. Datasets were generated using the equations implemented on a spreadsheet supplied by Dr AJ Olszowka (Model 1). This calculated exact values of arterial oxygen saturation for given values of inspired oxygen concentration. It represented the lungs as three compartments: a pure shunt and two ventilated regions, each with a homogeneous alveolar ventilation-perfusion ratio (V_A_/Q). The values of cardiac output, oxygen consumption, haemoglobin concentration, shunt fraction, and the distribution of blood flow and alveolar ventilation to the ventilated compartments can be set. The perfusion of one compartment whose V_A_/Q was reduced from 0.85 to 0.3 was set at 90% of non-shunt flow. A shunt was fixed at 2% and PIO2 was varied between 15 and 30 kPa, the SaO2 being derived in each case. Then with V_A_/Q held at 0.85 and shunt increased stepwise from 2-25 % the PIO2 was changed over the same range of PIO2 and SaO2 derived. The slope of the PIO2 vs SaO2 curve as it intersected the 21 kPa PIO2 line was calculated. To provide a consistency check of this analysis two patients with pre-existing lung disease and 12 hour continuous SpO2 monitoring from a previous study[3] had their PIO2 vs SpO2 plots reanalysed with a newly developed algorithm based on a three compartment lung,[5,6].

## RESULTS AND DISCUSSION

The left panels in Fig 2 show the effect on PIO2 vs SaO2 curves of reducing V_A_/Q or increasing shunt where the vertical line represents 21 kPa PIO2 breathing air at sea level. Reducing V_A_/Q from normal (circa 0.85) shifted the curve gradually to the right so that its gradient as it intercepted the 21kPa line became rapidly steeper. Furthermore, as V_A_/Q fell, the intercept of the curve on the 21kPa line showed SaO2 falling abruptly; red arrows indicate a 7% fall in SaO2 as V_A_/Q fell from 0.5 to 0.4 and a 24% reduction in SaO2 as V_A_/Q fell from 0.4 to 0.3. The panels on the right show the greater effect on gradient of reducing V_A_/Q than of increasing shunt. The SpO2 will become unstable once V_A_/Q has fallen to 0.5; at a V_A_/Q of 0.4 a ±1 kPa change in PIO2 gives an 8% change in SpO2 with the likelihood of profound hypoxemia with a further small fall in V_A_/Q. In contrast, with a 15% shunt, a ±1 kPa change in PIO2 gives <2% change in SpO2.

**Fig 2.**
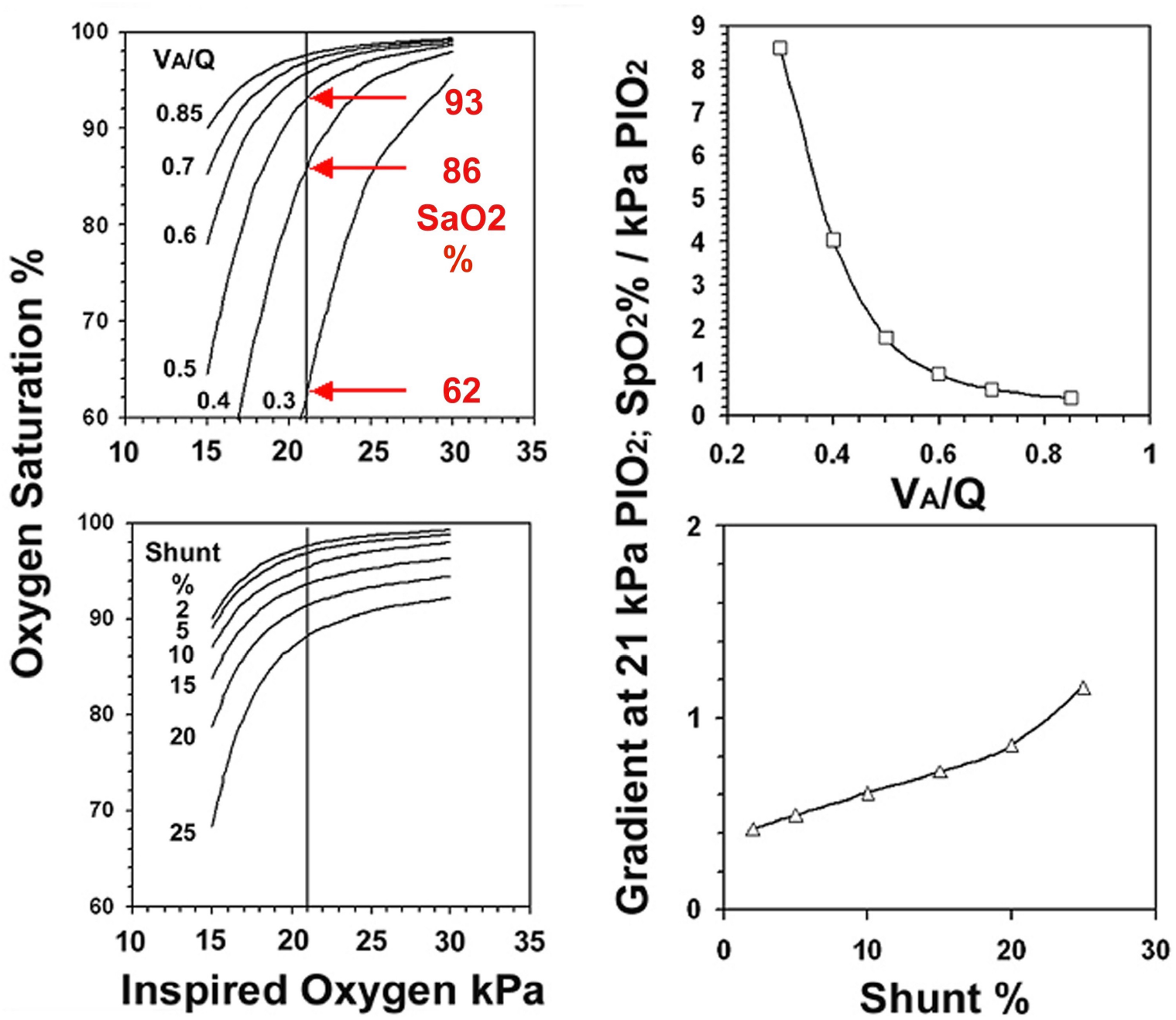
Effect of reducing VA/Q or increasing shunt on the PIO2 vs SaO2 curves (left panels) and on the gradient of these curves as they intersect the 21 kPa line (right panel). See text

To illustrate the clinical effect of V_A_/Q vs Shunt on SpO2 stability are two patients with a modest impairment of gas exchange, one with increased shunt the other with decreased V_A_/Q re-analysed from Fig11 Ref 4. (Fig 3). Left panels show oxygen dissociation (red) and normal lung PIO2 vs SpO2 curves (blue). Patients A and B had dark green PIO2 vs SpO2 curves derived by changing PIO2 stepwise and analysing the resulting PIO2 vs SpO2 data pairs with a new computer algorithm, [5,6]. The curve plateau was displaced downwards in A by a moderate shunt (14%) but not shifted to the right. When breathing air the gradient of curve A as it intercepted the 21kPa PIO2 line was 0.5 and similar to the normal lung. In B the whole curve was shifted to the right by the V_A_/Q reduced to 0.48. This patient had a shunt of 12% but the effect on SpO2 was outweighed by reduced V_A_/Q increasing curve gradient at the intercept to 2% SpO2 per kPa PIO2 which is similar to the result of the mathematical model in the right upper panel of Fig 2.

**Fig 3.**
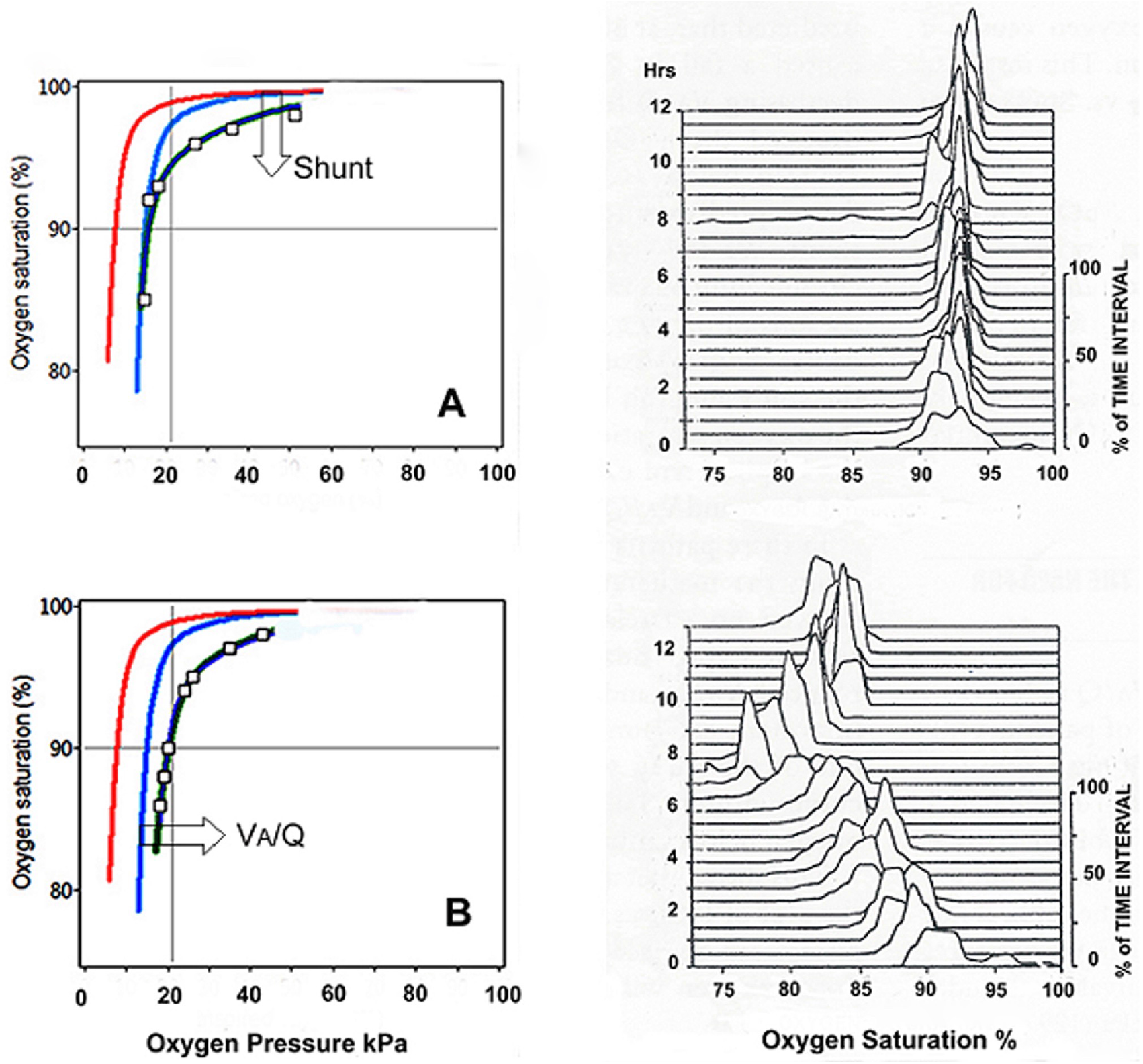
ODC (red) and PIO2 vs SpO2 curves for normal lungs (blue) compared to patients A & B (dk.green), left panels. Pt A curve was displaced downwards by a shunt and in Pt B shifted to the right by reduced VA/Q. Continuous SpO2 monitoring, breathing air, displayed with compressed spectral arrays. Pt A showed superimposed SpO2 peaks throughout the study whereas in B peaks were broad and unstable. Re-analysed and redrawn from Fig 11,[4].

Compressed spectral arrays (or Waterfall plots) were used to display the continuously measured SpO2 during the 12 hour monitoring periods breathing air. These showed the distribution pattern of SpO2 in 30 minute intervals, each moving up the page. Patient A with an increased shunt showed a stable pattern with superimposed peaks varying 1-2% around the median, closely resembling the normal lung pattern [4], but with a lower than normal median SpO2 circa 93-94%. In contrast Patient B with reduced V_A_/Q had broad peaks with SpO2 drifting downwards from >95 to 75% during the study. Large falls in SpO2 are consistent with small reductions in V_A/_Q below 0.5. Compared to the highly unstable SpO2 with a modest reductions of V_A_/Q to 0.5, it would be expected from Fig 2 that a patient with a 25% shunt breathing air would have a stable saturation down to 88%SpO2.

Instability of SpO2 breathing air with SpO2 varying >5% in 30 minutes is a sensitive marker of a reduction in V_A_/Q, e.g. to <0.5. The shape of the P_I_O_2_ vs SpO_2_ curve indicates more specifically the nature and magnitude of the underlying gas exchange abnormality; only a pencil and paper are required with PIO2 varied in three or more steps using a Ventimask,[10] In practice reduced V_A_/Q and and increased shunt often co-exist in different proportions giving complex effects on curve shape. A computer program plots the curve and generates a numerical estimate of V_A_/Q and Shunt from SpO_2_ vs P_I_O_2_ data pairs when P_I_O_2_ is changed stepwise,[5,6].

Other groups have reported on SpO2 monitoring of at risk Covid-19 patients in a domestic “virtual ward” setting. One chose a threshold for admission of 2% less than a 96% target, [11]. Another did not find such monitoring useful because of variability of implementation, [12] whereas a third reported on remote monitoring of 26 patients after discharge from hospital with Covid-19,[13]. In the latter group there were 5 “alerts” and 4 re-admissions with a median of 91% SpO2 and a lowest reading of 82%. None commented on unstable SpO2.

## CONCLUSION

A modest reduction in V_A_/Q to between 0.5 and 0.4 causes a right shift of the PIO2 vs SpO2 curve and a considerable increase in the gradient of this line as it intersects the 21kPa line. Small changes in ventilation and/or a further small reduction in V_A_/Q causes large changes in SpO2 circa >4%. While SpO2 instability breathing air is a sign of reduced of V_A_/Q it is not, per se, diagnostic of Covid-19 pneumonia. SpO2 varying substantially both below and above the 94% target threshold may be confusing to patients and their health teams. An at risk Covid-19 patient with previously normal SpO2 but develop ing unstable SpO2 with episodes of profound hypoxaemia out of proportion to radiological change may well be compatible with the reduced V_A_/Q of Covid-19 pneumonia.

## Data Availability

All data included in manuscript is available for review

## ACKNOWLEDGMENTS

The author thanks Dr A Olszowka, Department of Physiology, University of Buffalo, New York, USA for providing his pulmonary gas exchange program and Mr Jon Brassey, ***Trip Database***, for help in the preparation of this paper.

